# COVID-19 case forecasting model for Sri Lanka based on Stringency Index

**DOI:** 10.1101/2020.05.20.20103887

**Authors:** Achala U. Jayatilleke, Sanjeewa Dayarathne, Padmal de Silva, Pandula Siribaddana, Rushan A.B. Abeygunawardana, Olivia Nieveras, Nilanthi de Silva, Janaka de Silva

## Abstract

**Introduction:** Sri Lanka has been able to contain COVID-19 transmission through very stringent suppression measures implemented from the onset. The country had been under a strict lockdown since 20 March 2020, and currently the lockdown is being relaxed gradually. The objective of this paper is to describe a projection model for COVID-19 cases in Sri Lanka applied to different scenarios after lockout, utilizing the stringency index as a proxy of total government response.

**Methods:** COVID-19 Stringency Index (C19SI) is published and updated real time by a research group from Oxford university on 17 selected mitigation and suppression measures employed by different countries. We have mapped and validated the stringency index for Sri Lanka and subsequently validated the projection model in two phases. Predictions for the base-case scenario, less stringent scenarios, advanced relaxation scenarios, and high-risk districts were done with 95% confidence intervals (95%CI) using the validated model, utilizing data up to 10^th^ May.

**Results:** C19SI was able to accommodate all of the government responses. The model using validated C19SI could predict number of cases with 95% confidence for a period of two weeks. The model predicted base-case scenario of 815 (95%CI, 753–877) active cases by 17 May 2020 and 648 (95%CI, 599–696) cases for the high-risk districts by 18 May 2020. The model further predicted 3,159 (95%CI, 2,928–3,391) cases for 75% stringency and 928,824 (95%CI, 861,772–995,877) cases for 50% stringency. Advancing normalcy by three weeks resulted in the case load increasing to 4526 (95%CI, 4309–4744) by 30 June 2020.

**Conclusion:** The proposed prediction model based on C19SI provides policy makers an evidence based scientific method for identifying suppression measures that can be relaxed at the most appropriate time.

## Introduction

The World Health Organization (WHO) declared the COVID-19 outbreak as a pandemic on 11 February 2020^1^. The first case of Covid-19 was reported in Sri Lanka on 27 January 2020 in a 44-year-old Chinese woman from Hubei Province in China^2^. Almost six weeks later, Sri Lanka identified the first COVID-19 patient who contracted the disease locally^3^. The number of COVID-19 cases reported since then showed a gradual increase and included both imported and locally transmitted cases. Since the country followed an aggressive policy of contact tracing and isolation the health authorities were able to contain the situation^4–6^. However, on 22 April 2020, a sailor from a Sri Lanka Navy camp outside the capital, Colombo, who was actively engaged in contact tracing, tested positive for COVID-19, and 19 more sailors tested positive on the same day^7^. Since then the outbreak has escalated, but has been contained within Navy camps, and many of the COVID-19 positive sailors have been asymptomatic^8^. Sri Lanka has to date reported nine deaths attributable to COVID-19, with the latest reported death on 5 May 2020^9^.

Based on the four-stage classification of epidemics published by the WHO; ie. no cases, sporadic cases, community clusters and community transmission, Sri Lanka is currently in the category of community clusters^10^. At the time of writing this paper, Sri Lanka has been able to contain disease transmission, preventing the occurrence of ‘community transmission’, where the risk of health services in the country being overwhelmed is very high.

The measures countries take to tackle the effects of COVID-19 can be classified broadly as mitigation methods and suppression methods^11^. In mitigation, the focus is to slow down the spread of the virus and achieve a staggered onset. Although such measures would continue to allow the spread of the disease, the health services are unlikely to be overwhelmed due to the slow spread of the virus and communities would develop herd immunity, thereby easing the burden on the countries. The measures adopted by the Netherlands, Sweden and the United Kingdom (at least in the early stages of COVID-19 spread), were focused on mitigation^11,12^. Suppression methods were focused on stopping the spread completely, and the measures are generally more forceful than for disease mitigation. China, Vietnam and New Zealand adopted suppression measures which enabled them to flatten the curve and reduce the burden on their health services^13^. Some European countries such as Italy and Spain, changed their strategies from largely mitigation methods to suppression after the numbers of positive cases and deaths increased exponentially^11^. Both methods, mitigation and suppression, have their positive and negative health, economic and social consequences. The steps adopted in Sri Lanka in tackling the epidemic were largely suppressive, from the onset. These included, quarantining of all inbound passengers to the country, suspension of inbound flights, closing of schools and universities, implementing an island wide curfew, proactive contact tracing, quarantining and enforcing social distancing^14^.

The responses of governments to the pandemic, depending on the focus on mitigation or suppression, may impart different stringency levels. For instance, Scandinavian countries and the Netherlands have attempted to tackle the spread of COVID-19 by adopting less stringent mitigation policies than countries such as Vietnam and China^12,13^. Thomas Hale et al describe an index, based on the degree of stringency of government policies towards managing the COVID-19 pandemic^15^. The stringency index, developed by a group of researchers from the University of Oxford, considers multiple indicators including policies such as school closures, travel bans, and financial indicators such as fiscal and monetary measures. The index simply refers to the strictness of government policies in each country and is not necessarily indicative of how effective such policies are. The stringency index varies from country to country and to varying degrees with time, depending on the local disease situation, as was seen in Singapore with the emergence of new disease clusters among immigrant workers^15^.

We believe that the stringency index could be a useful parameter in modeling COVID-19 disease control, especially when countries have adopted policies which may be classified as suppressive. While adopting strict policies seem to dampen the spread of COVID-19 and limit an acute rise in the numbers, there is little evidence as to how the disease may behave as countries start to ease control policies. This paper describes a projection model for COVID-19 cases in Sri Lanka applied to different scenarios utilizing the stringency index as a proxy of the total government response. We aim to supplement already existing projection models for COVID-19 in Sri Lanka and identify the change in COVID-19 disease spread with a changing stringency index and epidemiological trends. This could help decision makers to visualize the potential effects of policy changes from the perspective of the stringency of such measures.

## Methods

For this analysis, we extracted publicly available data from the official website of the Epidemiology Unit of the Ministry of Health and Indigenous Medical Services, Sri Lanka^16^. and other websites^17–20^. We have not used any personally identifiable data for this analysis. We extracted data relevant to cases reported from 27 January 2020 to 10 May 2020. We did not include the first case reported in Sri Lanka in the analysis, as the second case was reported more than six weeks later and none of the cases reported subsequently were linked to the first case. Relevant stringency indexes were extracted from Oxford COVID-19 Government Response Tracker listing the COVID-19 Stringency Index (C19SI)^15^ developed by the Oxford University, United Kingdom. We extracted Government control measures from the Ministry of Health and other Government sources^14,16–20^. We have plotted daily new cases and mapped the Government response against the C19SI. We have mapped out the many interventions applied in Sri Lanka in responding to the COVID-19 pandemic, in relation to the areas of the C19SI as indicated by C1–C8 (Containment and Closure), E1–E4 (Economical Response), H1–H4 (Health System Response) and M1 (Miscellaneous). Further, we plotted C19SI and cumulative number of confirmed COVID-19 cases for seven selected countries with a view to validating the index for its applicability as a projection tool. Since different countries had initiated their response measures at different points in time, we plotted the C19SI from 14 days prior to the onset of the first case in each country, considering the incubation period for COVID-19.

We have applied the SIR (susceptible, infected and removed/recovered) model, given its reliability as a model applicable to large populations facilitating the predictions in disease outbreaks^21,22^. In the SIR model, Susceptible (*S*) individuals are those at risk of infection, infected (*I*) indicates individuals who are infected, and recovered or removed (*R*) includes those who may have developed immunity or died. Movement between these three categories is governed by β and γ which describe the “rate” of the infection and the infectious period, respectively^23^. The COVID-19 Stringency Index was used as a proxy to replace the parameter β. The per-capita recovery rate, γ, is usually difficult to influence, except through the introduction of vaccines^24^. Hence γ is highly unlikely to change over time during the current COVID-19 pandemic. In the absence of adequate local data due to the limited number of cases, we have considered 11 days as the recovery period to estimate the parameter in this study^25^. Using the SIR model and Ordinary Least Squares (OLS) technique, we projected the number of COVID19 cases that would present based on existing stringency measures in Sri Lanka up to 11 May 2020, and anticipated values for stringency index based on current arrangements for exit thereafter. Based on reported cases and the documented stringency index, the model was validated for its ability to predict the number of cases with 95% confidence for a period of 14 days. Since a new cluster was identified from the Welisara Navy Camp on 22 April 2020, which significantly changed the behavior of the epidemic, the validation was done based on two periods of time: (1) using data for 11 March to 8 April and projecting from 9 to 22 April, and (2) using data from 11 March to 26 April and projecting from 27 April to 10 May.

Once the model was validated, we refined the projection by applying the model to the entire country and high-risk areas respectively. Since S_0_ (the initial susceptible population) also plays a key role in estimating the number of possible inflected population, two approaches were adopted: (1) considering the entire population of Sri Lanka as susceptible (base-case scenario); (2) considering only the population in the high-risk districts of Colombo, Gampaha, Kalutara and Puttalam as susceptible. We used the midyear population for 2019 (21,203,000) and the population for individual districts estimated by the Department of Census and Statistics^26^ for this purpose.

Further, we estimated the number of cases that would have presented at hypothetical stringency index values of 75% and 50% (maintained up to 11 May 2020) and relaxed in a gradual manner. Finally, we extended the projection by advancing the dates of relaxing of stringency measures.

## Results

As shown in Figure 1, most of the initial cases reported in Sri Lanka were among expatriates who returned to the country from Italy, while the next significant cluster was observed among participants of a religious activity. After the 19^th^ of April, new cases were mainly due to two large clusters: one found within Colombo (the first infected person within the cluster had arrived from India); and the other at a Navy Camp housing more than 4,000 Navy personnel, most of whom were on active COVID-19 contact tracing duty. Figure 1 illustrates how Sri Lanka initiated its response to COVID-19 in advance of the first case and continued to heighten the response gradually between the first and second cases (6 weeks apart), and further escalated its response immediately after the second case, to a stringency index to 75% within one week and 97% within two weeks.

**Figure 1.**
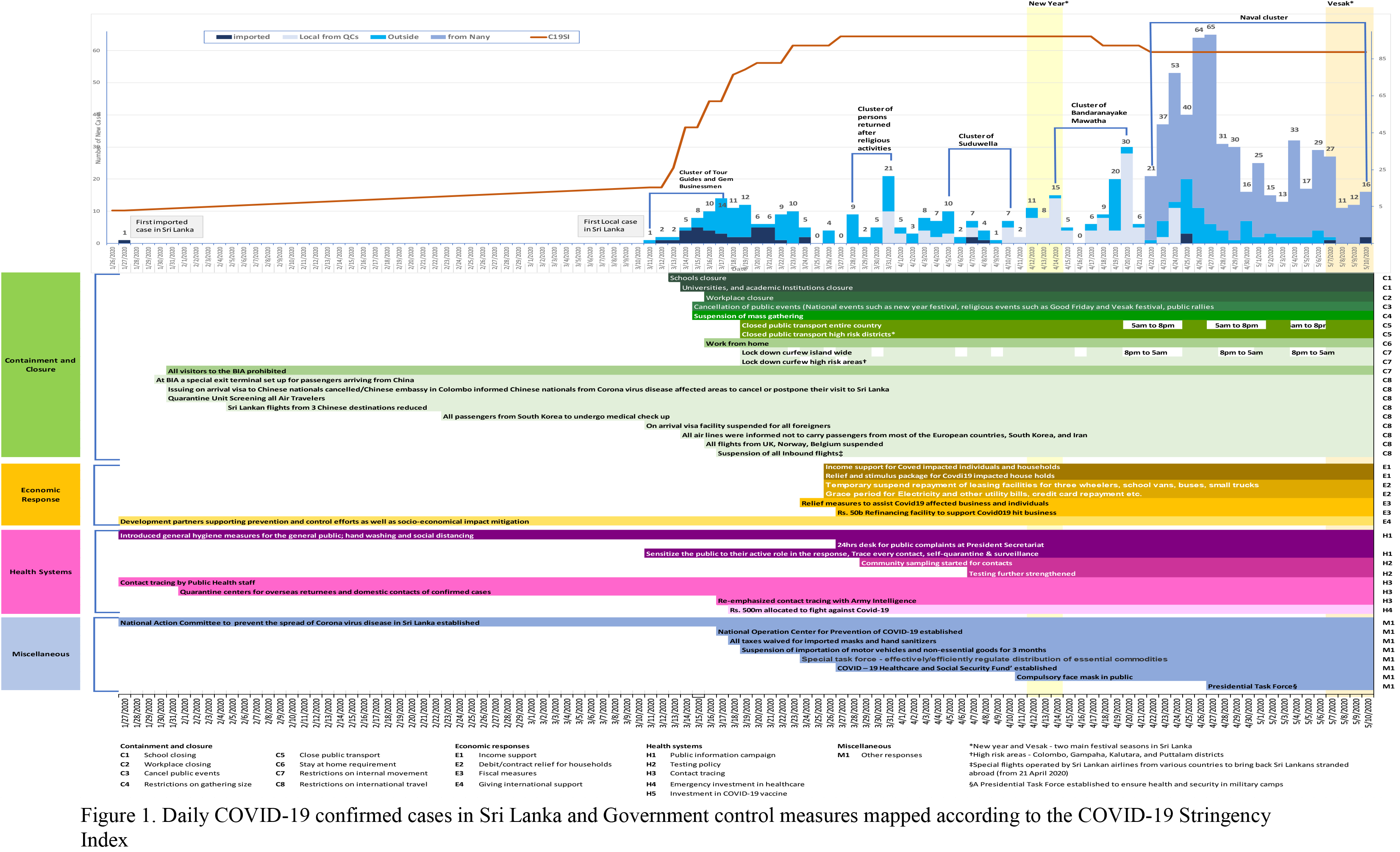
Daily COVID-19 confirmed cases in Sri Lanka and Government control measures mapped according to the COVID-19 Stringency Index

Figure 2 illustrates changes in C19SI for eight countries including Sri Lanka (Panel A), and the cumulative cases in each country over the same period of time (Panel B). Stringency Index for the countries were plotted from 14 days prior to the first reported case/s for each country. Compared to other countries, the Sri Lankan Government initiated control measures early. These measures closely resemble the measures taken by Vietnam. The control measures were escalated rapidly just after the first locally transmitted case was reported, and the Government Response Index reached its highest level within two weeks. Although Italy and Spain had escalations similar to that of Sri Lanka, it was after a considerable lag period. The United Kingdom and United States of America demonstrated an even longer lag period. Singapore and South Korea have gradually enforced stringency measures and demonstrate a stringency index below the other countries.

**Figure 2.**
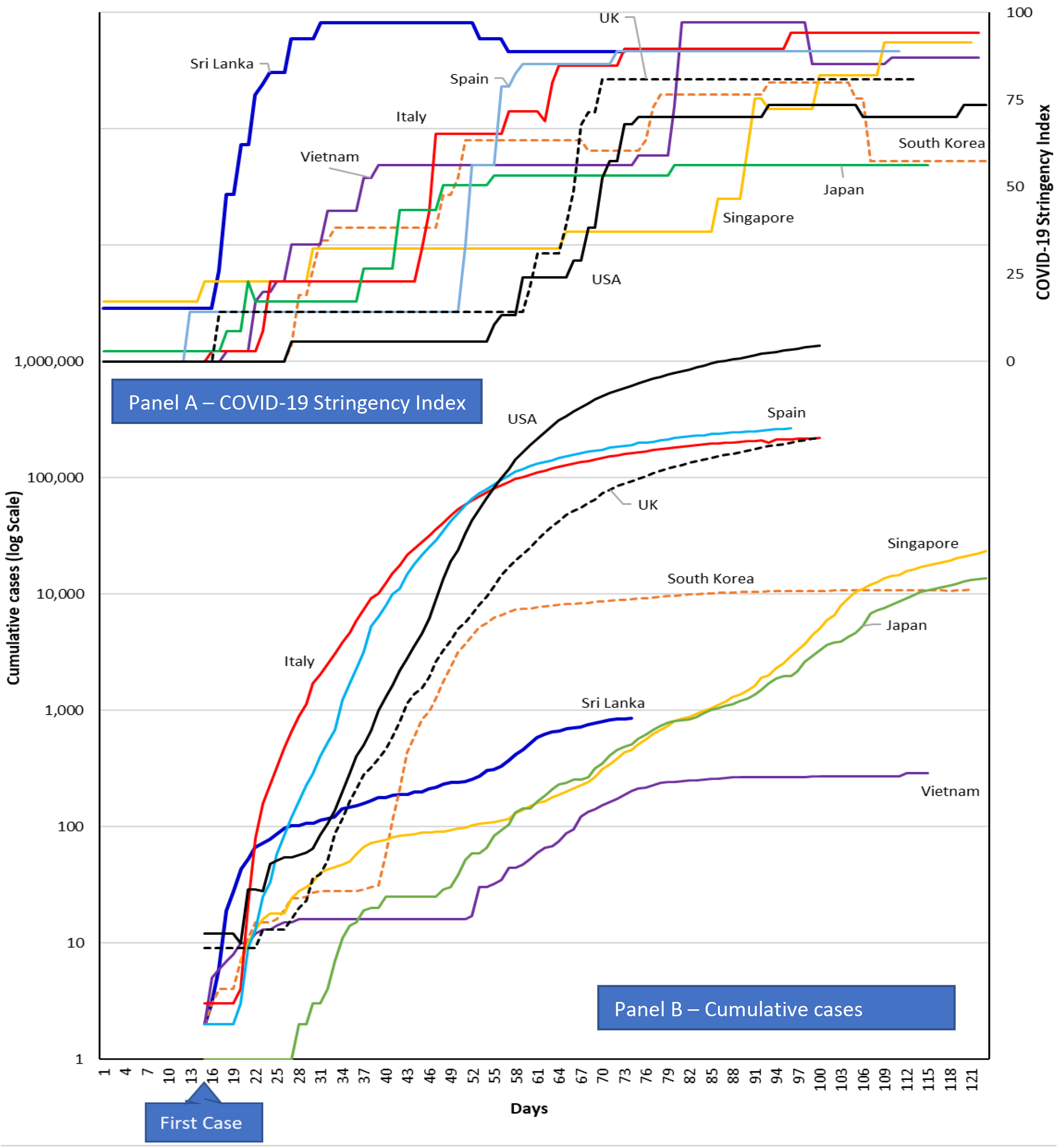
The behavior of COVID-19 Stringency Index for Sri Lanka and selected countries and cumulative cases^27^

Panel B of Figure 2, shows the number of COVID-19 cases reported for the same time period using a log scale. Countries that initiated their response early and escalated it before the case-load increased have been able to maintain a lower number of cumulative cases: Vietnam, Sri Lanka (less than 1000 cases) and South Korea (less than 10000 cases). Countries that had a high stringency index with a delayed escalation have demonstrated a larger number of cases: Italy, Spain, UK (around 200,000 cases) and USA (over 1.4 million cases).

Table 1 shows the projected number of new cases with 95% confidence interval for two-time periods: from 9 to 22 April and from 27 April to 10 May. For both phases, actual reported values lie within the 95% confidence interval of the prediction.

**Table 1.** Validation of the model

| Date | Predicted cases (95% Confidence Interval) | Actual active cases |
| --- | --- | --- |
| First phase based on 11 March to 8 April training data |  |  |
| 9-Apr-20 | 136 (120-151) | 133 |
| 10-Apr-20 | 139 (123-154) | 136 |
| 11-Apr-20 | 142 (126-157) | 137 |
| 12-Apr-20 | 145 (130-161) | 147 |
| 13-Apr-20 | 149 (133-164) | 152 |
| 14-Apr-20 | 152 (137-167) | 163 |
| 15-Apr-20 | 156 (140-171) | 166 |
| 16-Apr-20 | 159 (144-175) | 161 |
| 17-Apr-20 | 163 (148-178) | 160 |
| 18-Apr-20 | 171 (156-187) | 156 |
| 19-Apr-20 | 180 (165-196) | 167 |
| 20-Apr-20 | 189 (174-205) | 197 |
| 21-Apr-20 | 199 (183-215) | 201 |
| 22-Apr-20 | 214 (198-231) | 218 |
| Second phase based on 11 March to 26 April training data |  |  |
| 27-Apr-20 | 450 (418-482) | 447 |
| 28-Apr-20 | 470 (437-504) | 478 |
| 29-Apr-20 | 489 (454-523) | 503 |
| 30-Apr-20 | 505 (469-541) | 501 |
| 1-May-20 | 519 (482-556) | 511 |
| 2-May-20 | 531 (492-569) | 516 |
| 3-May-20 | 540 (501-579) | 524 |
| 4-May-20 | 546 (506-587) | 550 |
| 5-May-20 | 550 (509-591) | 547 |
| 6-May-20 | 551 (509-593) | 556 |
| 7-May-20 | 549 (506-591) | 575 |
| 8-May-20 | 544 (501-587) | 571 |
| 9-May-20 | 537 (493-580) | 517 |
| 10-May-20 | 526 (483-570) | 511 |

Figure 3 illustrates changes in the COVID-19 Stringency Index based on the following assumptions: Open government and private offices without limiting work but continue the lockdown for identified infection cluster locations in these areas till 1 of June 2020, further relaxation for social movement activities such as gradually opening of universities, removing traveling restrictions for the entire country by mid-June and “normalize” the country to operate without any quarantine centers and re-open schools mid-July. The results suggest that Sri Lanka will have 815 (95% CI, 753–877) active cases at the peak of the epidemic, occurring on 17 May 2020.

**Figure 3.**
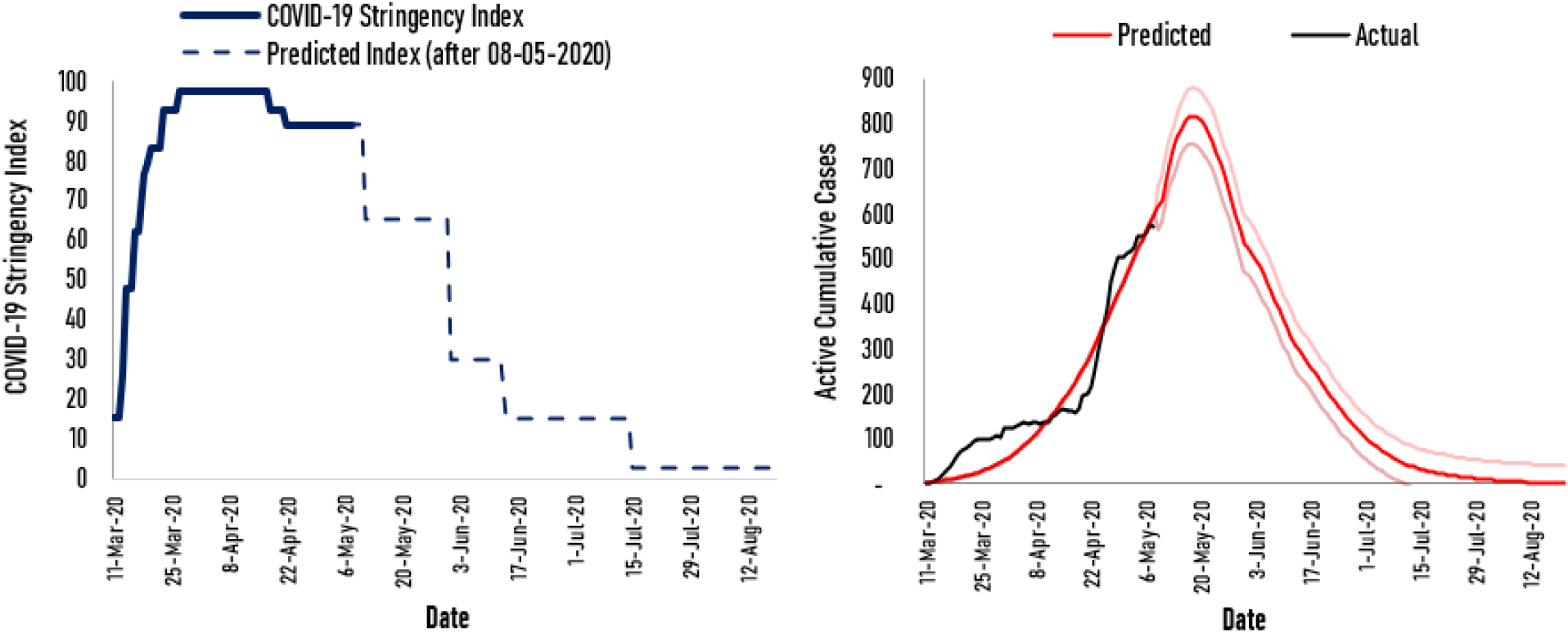
Changes in actual active cases and predicted active cases with β varying over time and fixed γ, –Base scenario

The projection was repeated out considering only the total population of four high-risk districts (Colombo, Gampaha, Kalutara, Puttalam) as susceptible, since the level of control measures implemented in the high-risk areas was more stringent than in other areas of the country. Therefore, the COVID-19 Stringency Index was modified to reflect the control measures implemented in these areas. This projection estimates that the number of active cases will peak by the 18 of May to 648 (95%CI, 699–696) for high-risk areas (Figure 4).

**Figure 4.**
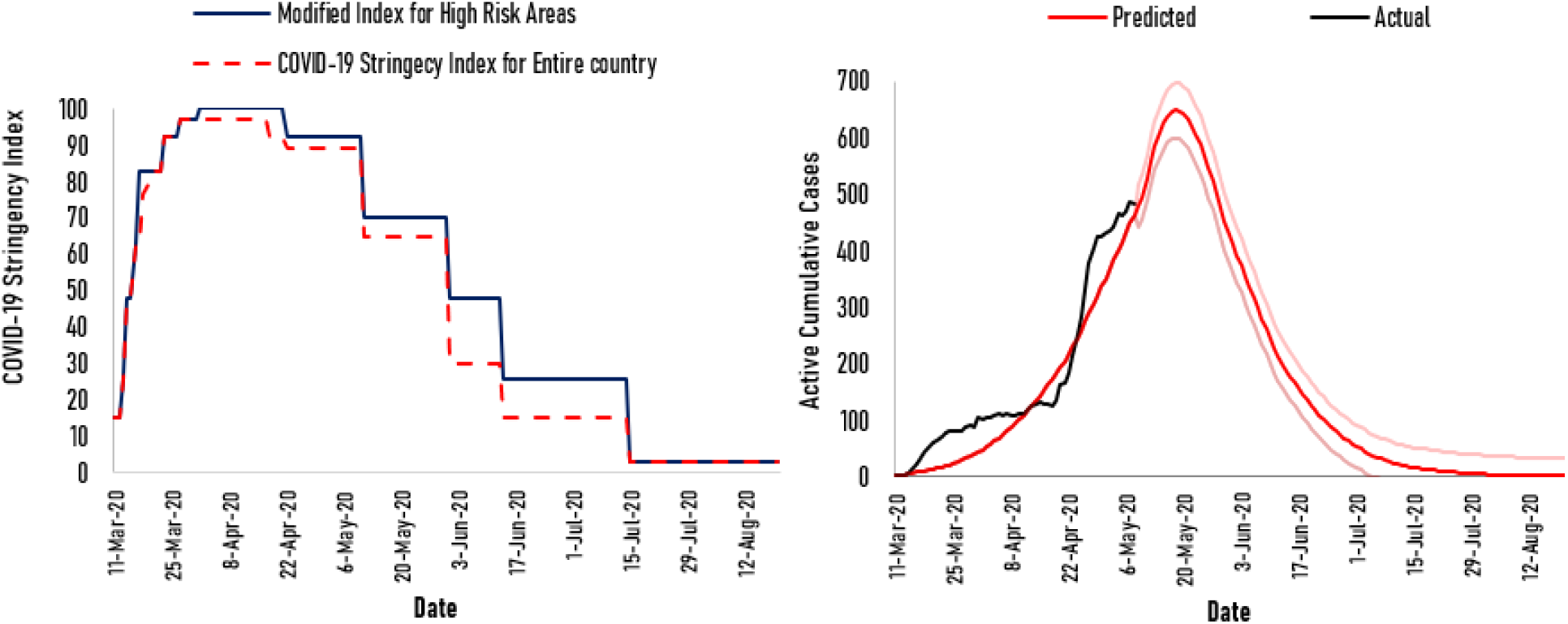
Changes in actual active cases and predicted active cases over time in high-risk areas.

As shown in Figure 5, we made projections for two other scenarios in addition to our base-case assumptions. Scenario 75% assumes only 75% of the actions under the base-case scenario will be implemented by the government. This would result in a peak of 3,159 cases (95%CI, 2,928–3,391) by 20 of May 2020. Scenario 50% assumes only 50% of the actions under the base-case scenario will be implemented by the government. This would result in a peak of 928,824 (95%CI, 861,772–996,877) cases by 25 of May 2020.

**Figure 5.**
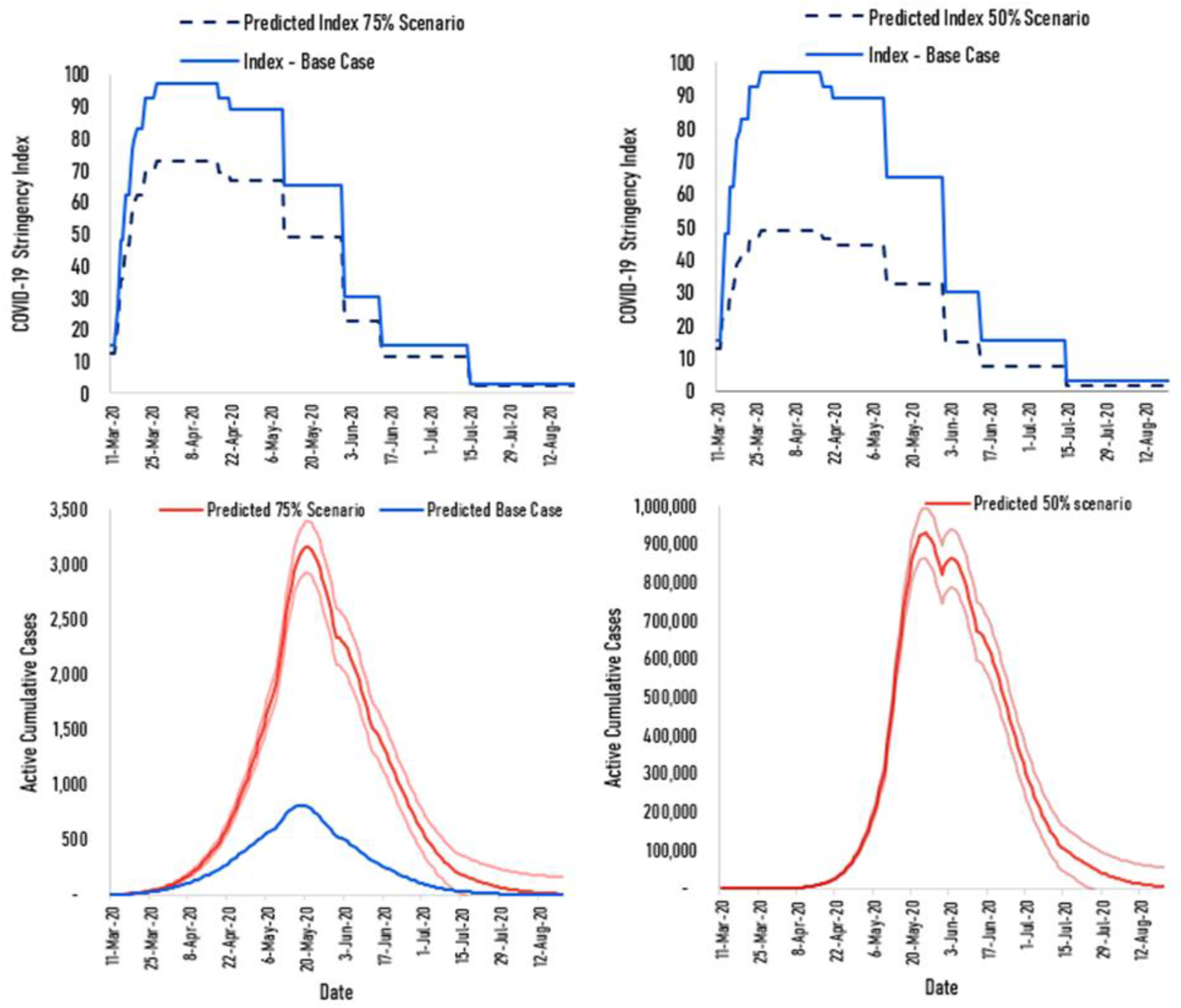
Changes in the Modified COVID-19 Stringency Index and predicted active cases for two scenarios; 75% scenario and 50% scenario

Figure 6 shows the possible impact of advancing the relaxation of each level of control by a week so that near normality would be advanced by one month. The model predicts a sharp increase in active cases after 13^th^ June where the peak number of active cases would rise to 4,526 (95%CI, 4,309–,4744) by 30^th^ of June 2020.

**Figure 6.**
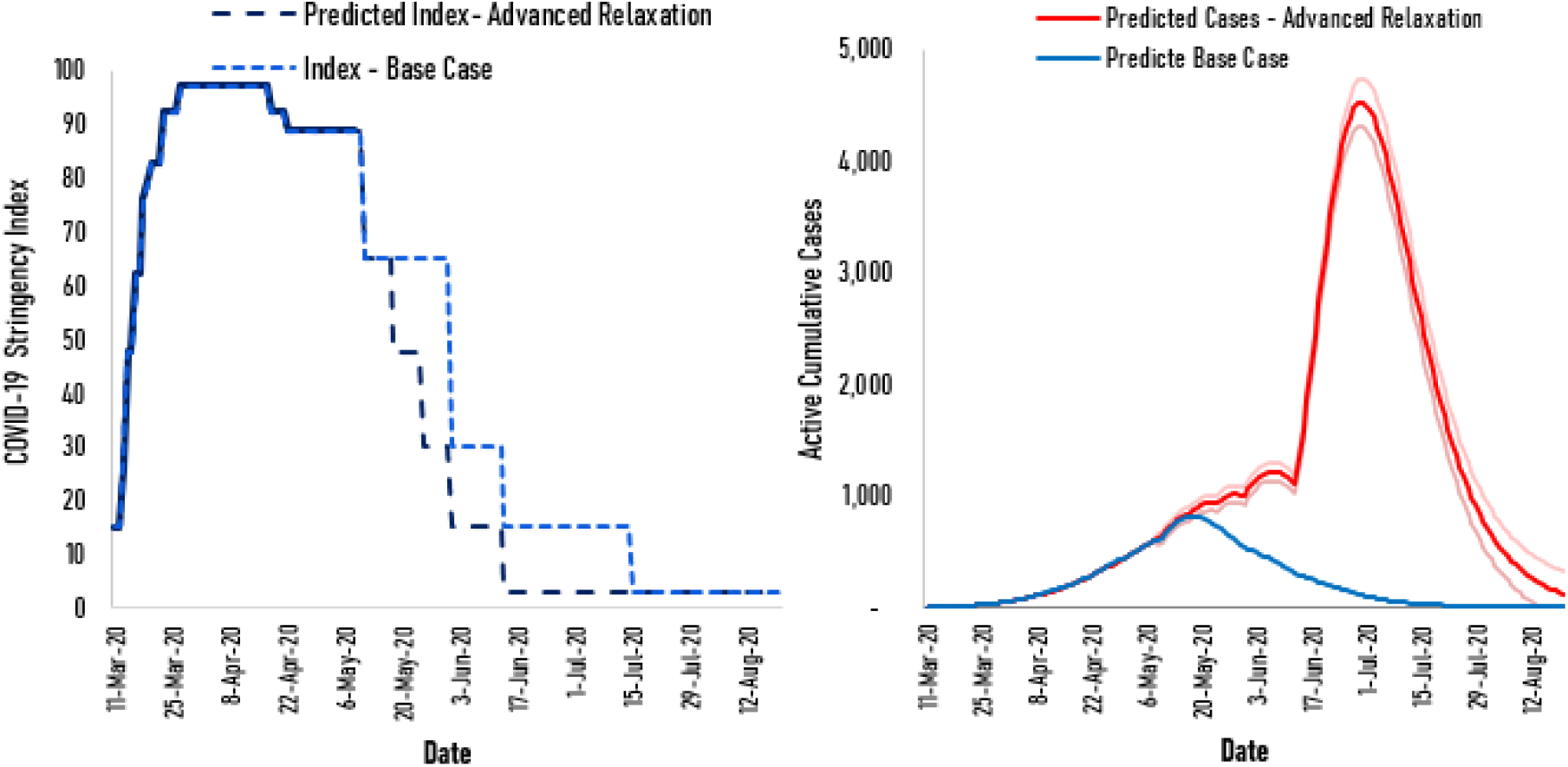
Advanced Relaxation scenario

## Discussion

In this paper, we have used the COVID-19 Stringency Index as a proxy to manipulate the parameter β in a SIR model, in order to enhance the predictability of future movements of the epidemic curve, as it is linked to practical actions. The methodology of estimating of C19SI is well documented in the BSG working papers. The extent of caseload is inversely related with the time taken to initiate control measures and the C19SI. The C19SI represents a composite indicator based on select response measures that have been globally practiced in suppressing the spread of COVID 19 and is updated regularly by the Oxford university research team. Therefore, we identified the C19SI as a proxy measure to be used as parameter β of SIR model. This model was able to predict the cases within 95% confidence for a period of two weeks. The second prediction had the full trend from 11 March to 26 April which included the cluster that emerged from the Navy camp which resulted in a three-fold increase in the number of cases for 4–5 days. Yet the prediction model run with the full set of values from 11 March to 26 April was able to predict accurately with 95% confidence the case load for two weeks starting 27 April. Therefore, the model had successfully factored in the surge of cases that emerged from the Navy cluster.

This would facilitate the real time estimation of the country specific C19SI for Sri Lanka, taking into consideration the response measures that are being considered for relaxation and projecting the case load based on the model using the C19SI at national and subnational levels. Based on the current pattern of cases and the suppression measures, in place the model predicted some 872 active cases at the peak of transmission. These predictions are in line with some other local predictions done by Weerasinghe (2020) using a SIR model with an active caseload of 1000 peaking after 90 days of onset^28^ and a separate group predicting based on a logistic regression model a total case load of 998 (+/− 6 cases) and ending by 20 July^19^. Similar findings are obtained from the model proposed by^29^ also using a SIR model. Further, based on our analysis, we have projected for C19SI or Stringency Index to be relaxed from 94 to 80 on 14 May and further relaxed to 60 by 1 June. The projection model has estimated that the number of active cases would increase from 815 (95%CI, 753–877) peaking on 17 May to 4,526 (95%CI, 4,309–4,744) peaking on 30 June 2020.

Based on the information available on COVID-19 cases and deaths reported from different countries, it is clear that varying levels of mitigation and suppression measures have been employed by different countries. The comparison of C19SI with the corresponding number of cases of COVID 19 reported by each country demonstrates a relationship between the timing of the response measures and the combination of the response measures put in place. In their report Walker et.al from the Imperial College group suggested that delays in implementing suppression strategies could lead to worse outcomes and fewer lives saved. For example, if suppression strategies are initiated when death rates are at 0.2 per 100 000 population per week and are sustained, this will save up to 38.7 million lives globally. Even if suppression strategies are initiated late when the death rates are as high as 1.6 per 100 000 population per week this could still save up to 30.7 million lives. The observation that there are more than 1.4 million cases for USA and more than 250,000 cases in UK, Spain, Italy and Russia, and far fewer numbers of cases in Vietnam, New Zealand and Sri Lanka can be explained by the different suppression strategies adopted by these countries^30^.

The accuracy of traditional forecasting models largely depends on the availability of data. In epidemics there may be no data at all in the beginning and with limited data early on in the epidemic, making predictions very uncertain. We have only begun to understand the nature of COVID-19 due to it being a novel infection and do not have the full information to fashion epidemiological predication models readily. Obtaining quality data on the numbers of cases, deaths and tests have been a challenge. Concerns regarding poor quality of data at the onset of any epidemic and problems in getting sharable data readily have been previously documented. Further, we are also limited by the paucity of real time data on implementation of the mitigation and suppression measures employed and to what extent these measures were adhered to by different population groups at national and subnational levels. Although we have mapped many interventions, there was no evidence to support that all of them were adhered to universally. Further, we felt that it would have been more effective to have a detailed index for the country which captures all the measures implemented by health authorities and other agencies. Such an index would be a refinement of the C19SI.

## Conclusions and Recommendations

The Oxford COVID-19 Government Response Tracker documents the stringency by which a government implements mitigation and suppression measures in controlling the spread of COVID-19. This stringency index accommodates all measures that governments across the globe have implemented and is therefore considered valid in the context of Sri Lanka as well. The C19SI provides a framework for generating a composite index based on many interventions, which in turn was used as a basis for the proposed model (parameter β) in this paper. The predictions made through the model therefore reflects on the number of cases that may result when control measures are relaxed at each point in time. The prediction model has demonstrated its ability to predict the number of cases for 14 days in advance with a 95% confidence. This prediction model based on the C19SI will be a useful tool for the timing and the extent of relaxing the many suppression and mitigation measures implemented by governments.

## Data Availability

Not applicable

## Disclaimer

The authors alone are responsible for the views expressed in this article and they do not necessarily represent the views, decisions or policies of the institutions with which they are affiliated

